# Characterization of disease course and remission in early seropositive rheumatoid arthritis: Results from the TACERA longitudinal cohort study

**DOI:** 10.1101/2020.03.08.20028142

**Authors:** RA-MAP Consortium, Brian Tom

**Author notes:** **Correspondence to:** Dr Brian Tom (PhD), MRC Biostatistics Unit, University of Cambridge, United Kingdom.

## Abstract

**Background:** To characterize disease course and remission in a longitudinal observational study of newly diagnosed, initially treatment naïve patients with seropositive rheumatoid arthritis (RA).

**Methods:** Patients with early untreated seropositive RA were recruited from 28 UK centres. Multiple clinical and laboratory measures were collected every 3 months for up to 18 months. Disease activity was measured using DAS28-CRP and SDAI. Logistic regression models examined clinical predictors of 6-month remission and latent class mixed models characterized disease course.

**Results:** We enrolled 275 patients of whom 267 met full eligibility and provided baseline data. According to SDAI definition, 24.3% attained 6-month remission. Lower baseline HAQ and SDAI predicted 6-month remission (p=0.002 and 0.021). Alcohol intake and baseline prescribing of methotrexate with a second DMARD (versus monotherapy without glucocorticoids) were also predictive. Three distinct SDAI trajectory subpopulations emerged; corresponding to an Inadequate Responder group (6.5%), and Higher and Lower Baseline Activity Responder groups (22.4% and 71.1%). Baseline HAQ and SF-36 MCS only distinguished these groups. Additionally a number of baseline clinical predictors correlated with disease activity severity within subpopulations. Beneficial effects of alcohol intake were found across subpopulations.

**Conclusions:** Three distinct disease trajectory subpopulations were identified. Differential effects of functional and mental well-being, alcohol consumption and baseline RA medication prescribing on disease activity severity were found across subpopulations. Heterogeneity across trajectories cannot be fully explained by baseline clinical predictors. Biological markers collected early in disease course (within 6 months) may help patient management and to better target existing and novel therapies.

## BACKGROUND

Early diagnosis and prompt, tight control of disease activity have been proven to alter long-term course of rheumatoid arthritis (RA) and limit structural damage and long-term disability.(1) In spite of a range of synthetic and biological disease modifying anti-rheumatic drugs, clinical remission is achieved and sustained in a minority and drug-free remission remains rare.(2) This may be due to many factors. For example, there are no validated instruments for reliably predicting prognosis. Nor is it possible to confidently predict which patients will respond more favourably to one particular drug or drug combination over another. Furthermore, our understanding of low disease activity states is limited.(3,4) The ability to employ the most appropriate ‘treat-to-target strategy’ to the right set of early RA patients would be a major advance. Recent evidence suggests that, for early RA patients following a treat-to-target strategy, distinct trajectories of disease activity over the first year exist.(5) Moreover, evidence from long-term observational cohorts has suggested that distinct trajectories linked to prognosis exist; which may point to distinct immunopathogenic subsets.(6) Even ACPA positive disease is heterogeneous in outcomes (e.g. radiological progression), further adding to arguments favouring a stratified medicine approach.(7,8)

To facilitate the goals of the RA MRC/ABPI (RA-MAP) Consortium,(9) a cohort of newly diagnosed seropositive RA patients – the ‘Towards A CurE for RA’ (TACERA) cohort – was established and followed frequently and deeply phenotyped. The aim of the current study was two-fold. Firstly, we aimed to determine if baseline clinical factors and RA-prescribed medications are associated with 6- month remission in seropositive early RA. However, an analysis at a single time point neither reflects longitudinal disease activity patterns nor what factors predict, for example, sustained remission, fluctuating disease activity course, or gradual versus rapid response. Therefore secondly, we aimed to (i) determine whether different types of disease activity trajectory sub-populations exist within this inception cohort; (ii) identify factors associated with longitudinal disease activity; and (iii) determine whether differences in trajectory types (if they exist) associate with disease outcomes, baseline RA-prescribed medication, smoking status and alcohol consumption. Identification of factors associated with disease activity and characterization of subgroups may improve patients’ treatment and management and identify the most suitable patients for recruitment into trials.

## METHODS

### Patients

We recruited newly diagnosed patients ≥18 years of age with symptom duration <12 months who fulfilled either the 1987 American College of Rheumatology (ACR) or 2010 European League Against Rheumatism (EULAR)/ACR classification criteria for RA.(10,11) Patients were required to be ACPA and/or RF positive and naïve to DMARDs or glucocorticoid therapy. Additionally, for patients recruited there needed to be a clear intention by the supervising rheumatologist to commence therapy with DMARDs. Patients were excluded if they had significant comorbidities or if pregnant or wishing to conceive. Participation in trials impacting on patient’s treatment, immune status or disease activity during the study period was not permitted. Ethical approval was authorised by the National Research Ethics Service London Central Committee (Reference number: 12/LO/0469). Informed, written consent was obtained.

### Study Design

Subjects were recruited from 28 UK centres. Study sample size was determined as described in supplementary material. Following enrolment, subjects received treatment using conventional synthetic DMARDs (csDMARDs), with therapy adjustments made at the supervising rheumatologist’s discretion according to the National Institute for Health and Clinical Excellence (NICE) guidelines for RA management in adults.(12) Patients were followed for a period of 18, 12 or 6 months, dependent on time of enrolment (I.e. pre 01/07/14, 01/07/14-31/12/14, post 31/12/14), and seen quarterly for scheduled assessments. Clinical, laboratory, lifestyle, medication, patient-reported outcome measures (PROMs), extra-articular RA features, adverse events and biological samples were collected at visits. Additionally, radiographs of the hands and feet were performed at baseline and 12 months or baseline and 6 months for subjects entering within the third enrolment period.

### Biological Treatment

Subjects with inadequate clinical responses to csDMARDs and persistent high disease activity (DAS28 > 5.1) at 6 months could receive biological DMARDs (bDMARDs), according to NICE guidelines.

### Outcome measures

The co-primary outcomes were disease remission at 6 months and repeated disease activity over time. Disease activity was measured using both composite DAS28-CRP (4-component) and SDAI scores.(13–16) Disease remission was defined using both DAS28-CRP remission criterion (DAS28-CRP < 2.6) and the more stringent SDAI criterion (SDAI ≤ 3.3).

Secondary outcome measures included the ACR/EULAR Boolean definition of remission(17), DAS28- ESR, annualized rate of radiographic damage progression as measured by modified Larsen’s score(18–20), functional disability as measured by the Health Assessment Questionnaire (HAQ)(21), and quality of life using both the Medical Outcomes Study Short Form 36 Health Survey (SF-36) and EuroQoL five dimensions questionnaire (EQ-5D).(22–24)

### Statistical methods

Baseline and 6-month information were summarised using the mean, with accompanying standard deviation (SD) for continuous variables, whilst binary or categorical variables were summarised using frequency and percentage. Stepwise logistic regression models were employed to identify baseline predictors of 6-month clinical remission. Considered predictors comprised age, sex, ethnicity, body mass index, symptom duration, smoking status, baseline disease activity, HAQ, SF-36 Mental Component Score, alcohol consumption, serology (rheumatoid factor (RF) and ACPA), smoking, erosions and prescribing of (or baseline intention to prescribe) RA medications.

Latent class mixed models (LCMM) were used to cluster disease activity trajectories that may identify clinically important sub-populations and characterize disease activity longitudinally. Within each latent class, fixed and random patient-level intercepts and piecewise linear time effects (at 5 months) were considered for the linear mixed model part along with potential predictors. No covariates were included in the class membership model part. Latent class mixed models are likelihood-based methods, which are valid using only observed data, under a missing-at-random assumption. Associations of latent trajectory classes with outcomes, baseline RA medication prescribing, alcohol and smoking status were assessed either using analysis of variance or Fisher’s exact test. For the purpose of these association tests, patients were allocated to particular latent trajectory class where their calculated assignment posterior probability was highest.

Statistical analyses were performed using R statistical software.(25) Two patients, not prescribed RA medications within the first three months, were excluded from the regression analyses.

## RESULTS

### Patient Characteristics

Two hundred and seventy five patients were recruited of whom 270 fulfilled all eligibility criteria. Two eligible patients withdrew at baseline without providing any clinical information. A further patient who withdrew at baseline had some clinical information but insufficient to calculate disease activity scores. Table 1 summarises the baseline characteristics of the remaining 267 patients.

**Table 1:**
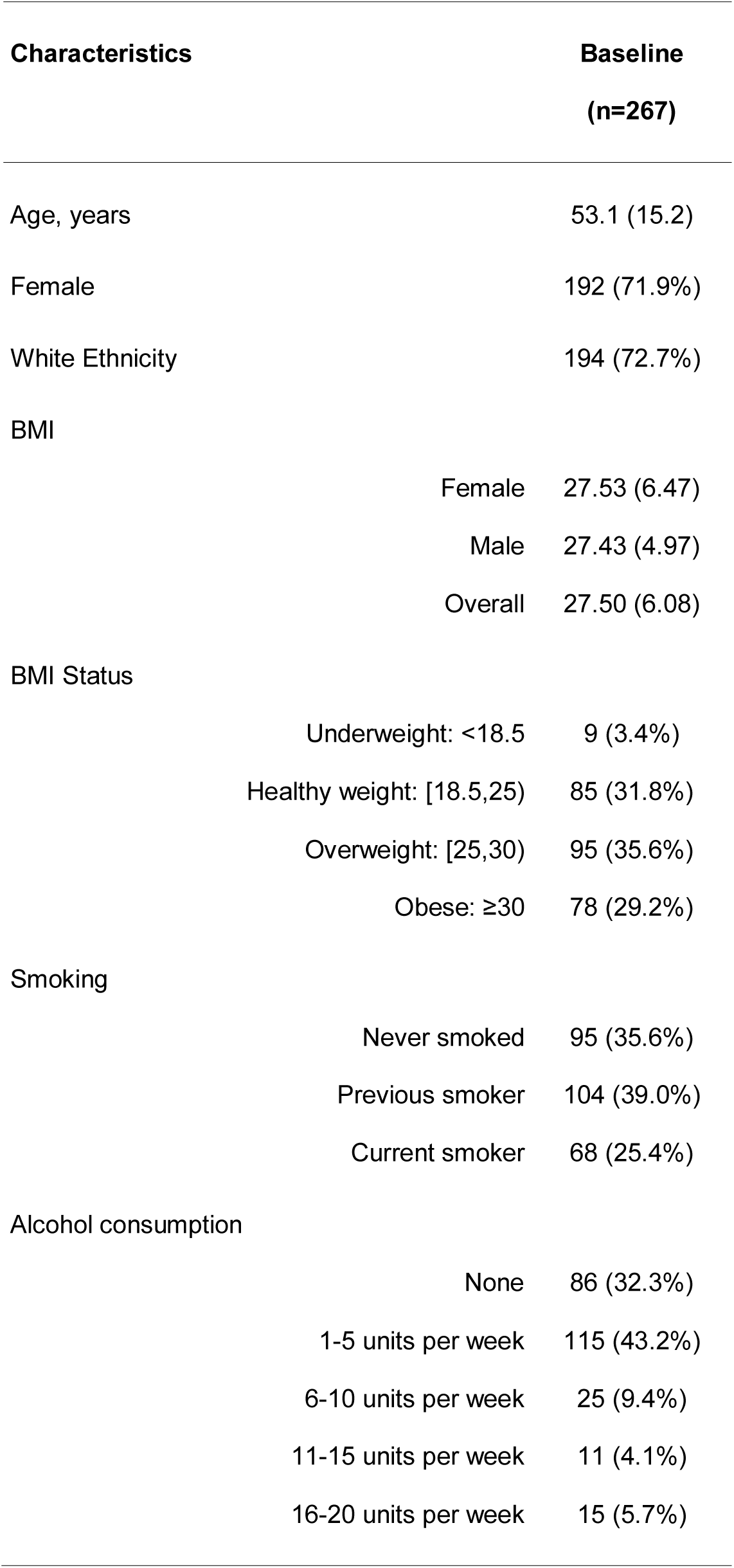
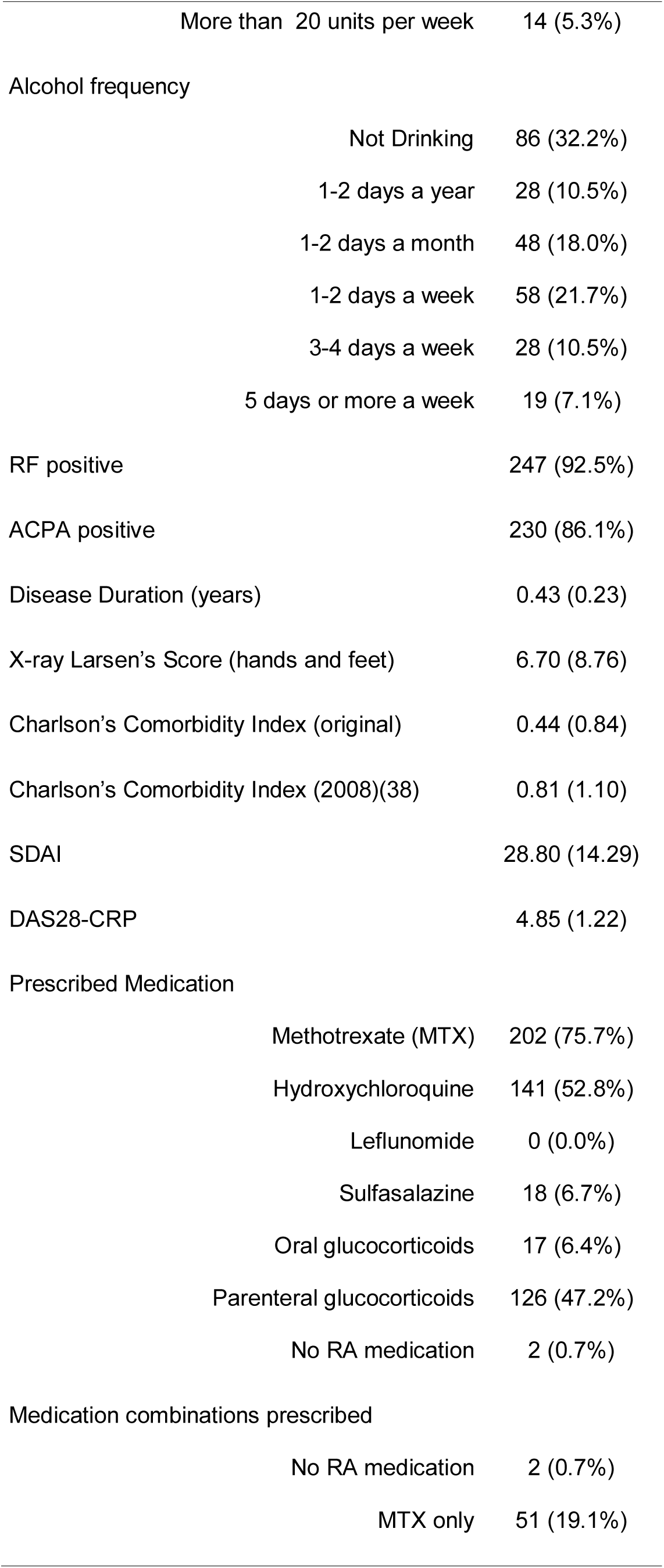
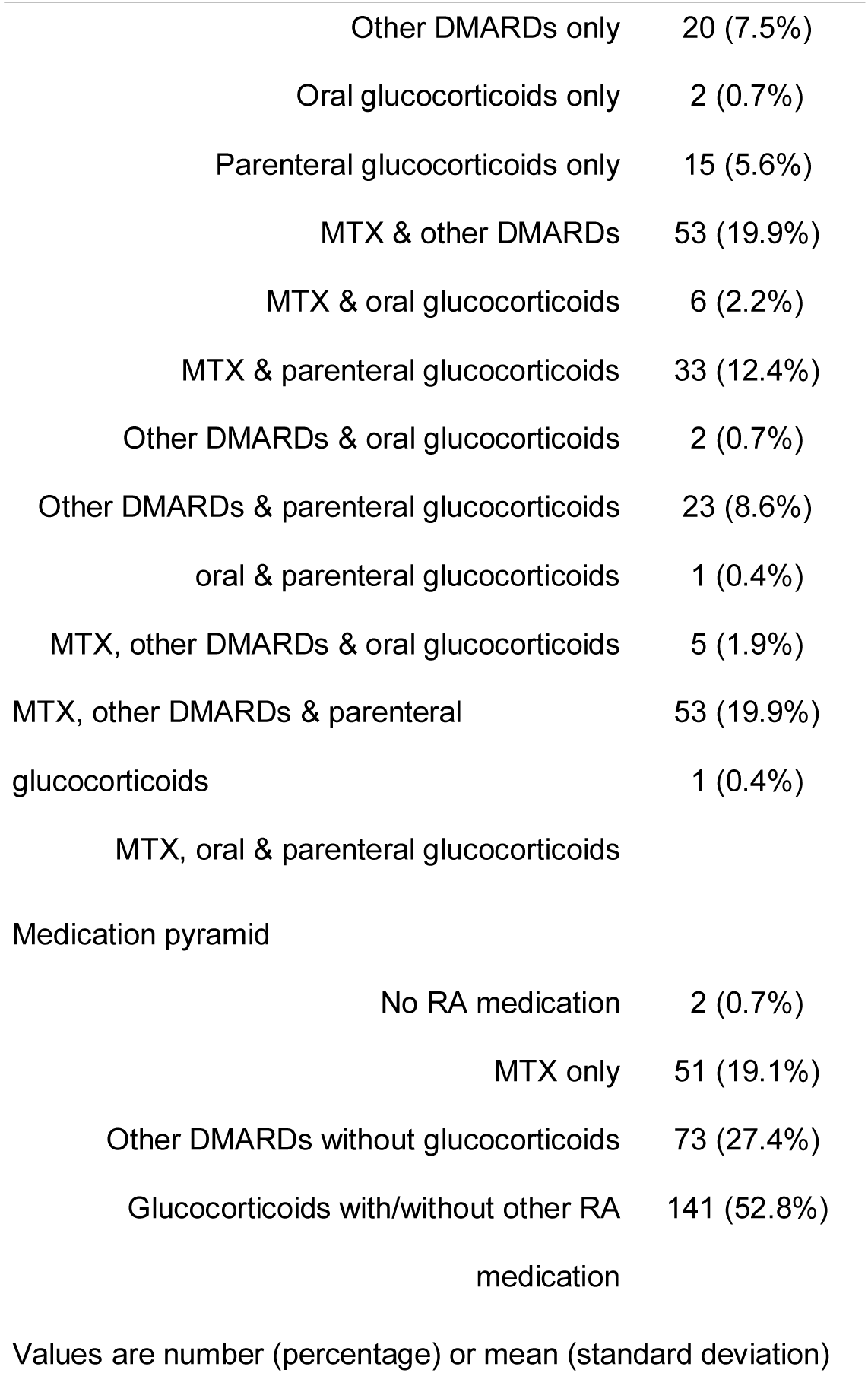
Baseline characteristics of patients in the TACERA study

| **Characteristics** |  | **Baseline**<br>**(n=267)** |
| --- | --- | --- |
| Age, years |  | 53.1 (15.2) |
| Female |  | 192 (71.9%) |
| White Ethnicity |  | 194 (72.7%) |
| BMI |  |  |
|  | Female | 27.53 (6.47) |
|  | Male | 27.43 (4.97) |
|  | Overall | 27.50 (6.08) |
| BMI Status |  |  |
|  | Underweight: <18.5 | 9 (3.4%) |
|  | Healthy weight: [18.5,25) | 85 (31.8%) |
|  | Overweight: [25,30) | 95 (35.6%) |
|  | Obese: ≥30 | 78 (29.2%) |
| Smoking |  |  |
|  | Never smoked | 95 (35.6%) |
|  | Previous smoker | 104 (39.0%) |
|  | Current smoker | 68 (25.4%) |
| Alcohol consumption |  |  |
|  | None | 86 (32.3%) |
|  | 1-5 units per week | 115 (43.2%) |
|  | 6-10 units per week | 25 (9.4%) |
|  | 11-15 units per week | 11 (4.1%) |
|  | 16-20 units per week | 15 (5.7%) |

|  |  |
| --- | --- |
| More than 20 units per week | 14 (5.3%) |
| Alcohol frequency |  |
| Not Drinking | 86 (32.2%) |
| 1-2 days a year | 28 (10.5%) |
| 1-2 days a month | 48 (18.0%) |
| 1-2 days a week | 58 (21.7%) |
| 3-4 days a week | 28 (10.5%) |
| 5 days or more a week | 19 (7.1%) |
| RF positive | 247 (92.5%) |
| ACPA positive | 230 (86.1%) |
| Disease Duration (years) | 0.43 (0.23) |
| X-ray Larsen's Score (hands and feet) | 6.70 (8.76) |
| Charlson's Comorbidity Index (original) | 0.44 (0.84) |
| Charlson's Comorbidity Index (2008)(38) | 0.81 (1.10) |
| SDAI | 28.80 (14.29) |
| DAS28-CRP | 4.85 (1.22) |
| Prescribed Medication |  |
| Methotrexate (MTX) | 202 (75.7%) |
| Hydroxychloroquine | 141 (52.8%) |
| Leflunomide | 0 (0.0%) |
| Sulfasalazine | 18 (6.7%) |
| Oral glucocorticoids | 17 (6.4%) |
| Parenteral glucocorticoids | 126 (47.2%) |
| No RA medication | 2 (0.7%) |
| Medication combinations prescribed |  |
| No RA medication | 2 (0.7%) |
| MTX only | 51 (19.1%) |
| Other DMARDs only | 20 (7.5%) |
| Oral glucocorticoids only | 2 (0.7%) |
| Parenteral glucocorticoids only | 15 (5.6%) |
| MTX & other DMARDs | 53 (19.9%) |
| MTX & oral glucocorticoids | 6 (2.2%) |
| MTX & parenteral glucocorticoids | 33 (12.4%) |
| Other DMARDs & oral glucocorticoids | 2 (0.7%) |
| Other DMARDs & parenteral glucocorticoids | 23 (8.6%) |
| oral & parenteral glucocorticoids | 1 (0.4%) |
| MTX, other DMARDs & oral glucocorticoids | 5 (1.9%) |
| MTX, other DMARDs & parenteral glucocorticoids | 53 (19.9%) |
| MTX, oral & parenteral glucocorticoids | 1 (0.4%) |
| Medication pyramid |  |
| No RA medication | 2 (0.7%) |
| MTX only | 51 (19.1%) |
| Other DMARDs without glucocorticoids | 73 (27.4%) |
| Glucocorticoids with/without other RA medication | 141 (52.8%) |

Values are number (percentage) or mean (standard deviation)

Briefly, the mean age at entry was 53.1 (SD of 15.2); 72% were female, 72.7% white, 31.8% healthy weight and the mean Charlson’s comorbidity index (CCI),(26) modified to exclude rheumatic disease as this applied to all subjects, was 0.44 (SD of 0.84), conforming to the intention of not recruiting patients with significant comorbidities.

Of the 267 patients, 130 (48.7%), 67 (25.1%) and 70 (26.2%) were enrolled in the first, second and third recruitment periods respectively. No statistically significant differences were found across these three groups with respect to disease-related variables, prescribed medication, and PROMs at baseline. However there were statistically significant differences found in the age, ethnicity, alcohol consumption and CCI distributions, with the third group being the oldest on average (51.5 vs 53 vs 56 years old), having the highest proportion of whites (71.5% vs 62.7% vs 84.3%), lowest proportion not consuming alcohol at entry (32.3% vs 44.8% vs 20%) and lowest levels of comorbidities (CCIs of 0.52 v 0.49 v 0.26).

Overall, patients had moderate to severe disease at baseline as measured by both DAS28-CRP and SDAI. After baseline assessment, MTX was prescribed to 75.7% of patients; 58.4% were prescribed non-methotrexate major DMARDs (51.7% hydroxychloroquine alone, 5.7% sulfasalazine alone and 1.1% both hydroxychloroquine and sulfasalazine); 6.4% received oral glucocorticoids (average prednisolone dose of 11.1 mg/day, range 4mg/day to 30mg/day) and 47.2% parenteral glucocorticoids (i.e. intra-articular, intravenous or intramuscular glucocorticoid administration). Overall, 33% were prescribed only one class/type of medication, whilst 66.3% were prescribed combination therapy (including with glucocorticoids) at baseline. Based on medication patterns at baseline, 26.6%, 19.9% and 52.8% were prescribed single RA therapy excluding glucocorticoids, dual therapy (i.e. MTX with another DMARD excluding glucocorticoids), and therapies that included glucocorticoid usage respectively. Initial pattern of therapy did not appear to associate with either baseline X-ray scores or CCI (p=0.56 and 0.23 respectively) but, as expected, was associated with disease activity measured using both SDAI and DAS28-CRP (p=0.01 and 0.004 respectively). Two subjects did not receive any RA medication by the time of their first follow-up assessment and were excluded from analyses.

### Disease activity, response and remission at 6 months

Two hundred and forty five patients (of the 267) were followed up to or beyond their 6-month assessment visit, with 75, 2, 60, 3 and 105 having their last assessment visit at 6, 9, 12, 15 and 18 months respectively. Of those recruited in the first, second and third periods, 80.8% (105), 83.6% (56) and 92.9% (65) reached their target follow-up assessment visits of 18 months, 12 months and 6 months respectively. Of the 245 followed up to or beyond 6 months, 239 attended their 6-month assessment visit. The disease activity and disease-related outcome measures for these 239 patients are summarized in Table 2. At 6 months, the mean (SD) DAS28-CRP and SDAI were 3.04 (1.25) and 11.37 (10.71) respectively (Supplementary Table 1). Based on EULAR response criterion, 110 (47.2%) patients had good response, 79 (33.9%) moderate and 44 (18.9%) no response. Regarding remission by different criteria, 97 (41.3%) patients achieved DAS28-CRP remission, 57 (24.3%) SDAI remission and 51 (21.5%) met the ACR/EULAR Boolean remission criteria reflecting increased stringency of definitions. All 57 patients in SDAI remission were in DAS28-CRP remission. There was excellent agreement between SDAI and ACR/EULAR Boolean remissions (Cohen’s kappa of 0.9).

**Table 2:**
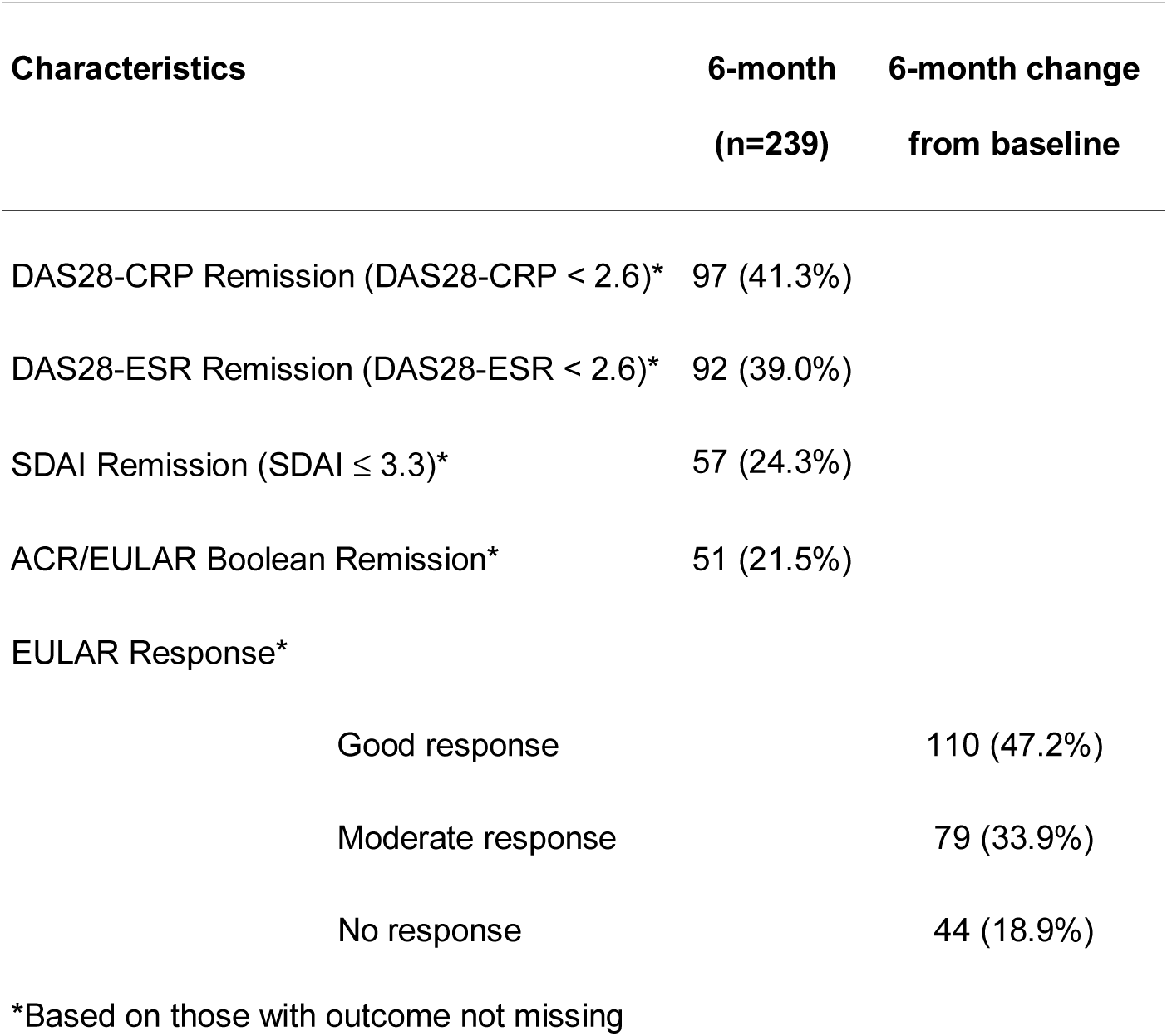
Disease activity response and remission at 6 months

| Characteristics | 6-month<br>(n=239) | 6-month change<br>from baseline |
| --- | --- | --- |
| DAS28-CRP Remission (DAS28-CRP < 2.6)* | 97 (41.3%) |  |
| DAS28-ESR Remission (DAS28-ESR < 2.6)* | 92 (39.0%) |  |
| SDAI Remission (SDAI ≤ 3.3)* | 57 (24.3%) |  |
| ACR/EULAR Boolean Remission* | 51 (21.5%) |  |
| EULAR Response* |  |  |
| Good response |  | 110 (47.2%) |
| Moderate response |  | 79 (33.9%) |
| No response |  | 44 (18.9%) |

\*Based on those with outcome not missing

Other outcome measures are described in Supplementary Table 1.

### Predictors of clinical remission at 6 months

Multivariate logistic regression models for SDAI and DAS28-CRP remissions are shown in Table 3. In both models, 6-month clinical remission was predicted by lower functional disability and disease activity at baseline. The odds ratios related to level of disease activity at baseline are 0.68 (95% CI: 0.49-0.95) for a 10 unit change in SDAI or 0.70 (95% CI: 0.52-0.94) for a 1 unit change in DAS28-CRP, reflecting that patients with a baseline disease activity 10 (or 1) units higher have approximately a thirty percent reduction in odds of achieving 6-month remission, controlling for other variables.

**Table 3:** Multivariate Logistic Regression Models for SDAI and DAS28-CRP remission at 6 months

| Predictors | Log (Odds Ratio) | Standard Error | p-value |
| --- | --- | --- | --- |
| **SDAI remission at 6 months** |  |  |  |
| Intercept | 0.128 | 0.547 | 0.814 |
| SDAI at Baseline | -0.038 | 0.017 | 0.021 |
| HAQ at Baseline | -0.979 | 0.313 | 0.002 |
| Alcohol consumption |  |  |  |
| 1-5 units per week v None | 1.278 | 0.461 | 0.006 |
| >5 units per week v None | 0.833 | 0.500 | 0.096 |
| Prescribed medication at baseline* |  |  |  |
| MTX and other DMARDs without glucocorticoids v therapies with glucocorticoids | 0.416 | 0.431 | 0.334 |
| monotherapy, not glucocorticoids v therapies with glucocorticoids | -0.724 | 0.433 | 0.094 |
| **DAS28-CRP remission at 6 months** |  |  |  |
| Intercept | 1.519 | 0.677 | 0.025 |
| DAS28-CRP at Baseline | -0.362 | 0.153 | 0.018 |
| HAQ at Baseline | -0.758 | 0.268 | 0.005 |
| Alcohol consumption |  |  |  |
| 1-5 units per week v None | 0.865 | 0.360 | 0.016 |
| >5 units per week v None | 1.367 | 0.414 | 0.001 |

\* The contrast of MTX and other DMARDs without glucocorticoids versus monotherapy gave a Log (Odds Ratio) estimate of 1.140 with standard error of 0.510 and a p-value of 0.026

Higher baseline functional disability was associated with a reduced likelihood of achieving SDAI remission (OR=0.88, 95% CI: 0.82-0.96 for a 0.125 increase in HAQ) or DAS28-CRP remission (OR=0.91, 95% CI: 0.85-0.97 for a 0.125 increase in HAQ) at 6 months.

Although both models suggested alcohol consumption increased the odds of remission at 6 months compared to not consuming alcohol, they gave conflicting ordering of effect sizes across the alcohol consumption categories of 1-5 units and greater than 5 units per week. Moreover, the SDAI model included effects of baseline prescribed medication, with evidence that being prescribed dual combination of MTX and a second DMARD increased the likelihood of SDAI remission compared to receiving monotherapy (OR 3.13; 95%CI 1.15-8.50; p=0.026).

### Characterising disease activity over time

Figure 1 (left hand side) shows the observed individual trajectories of SDAI and DAS28-CRP for the 267 patients. From the figure there is evidence of substantive within- and between-patient variation in disease activity profiles and potential distinct trajectory subtypes.

**Figure 1:**
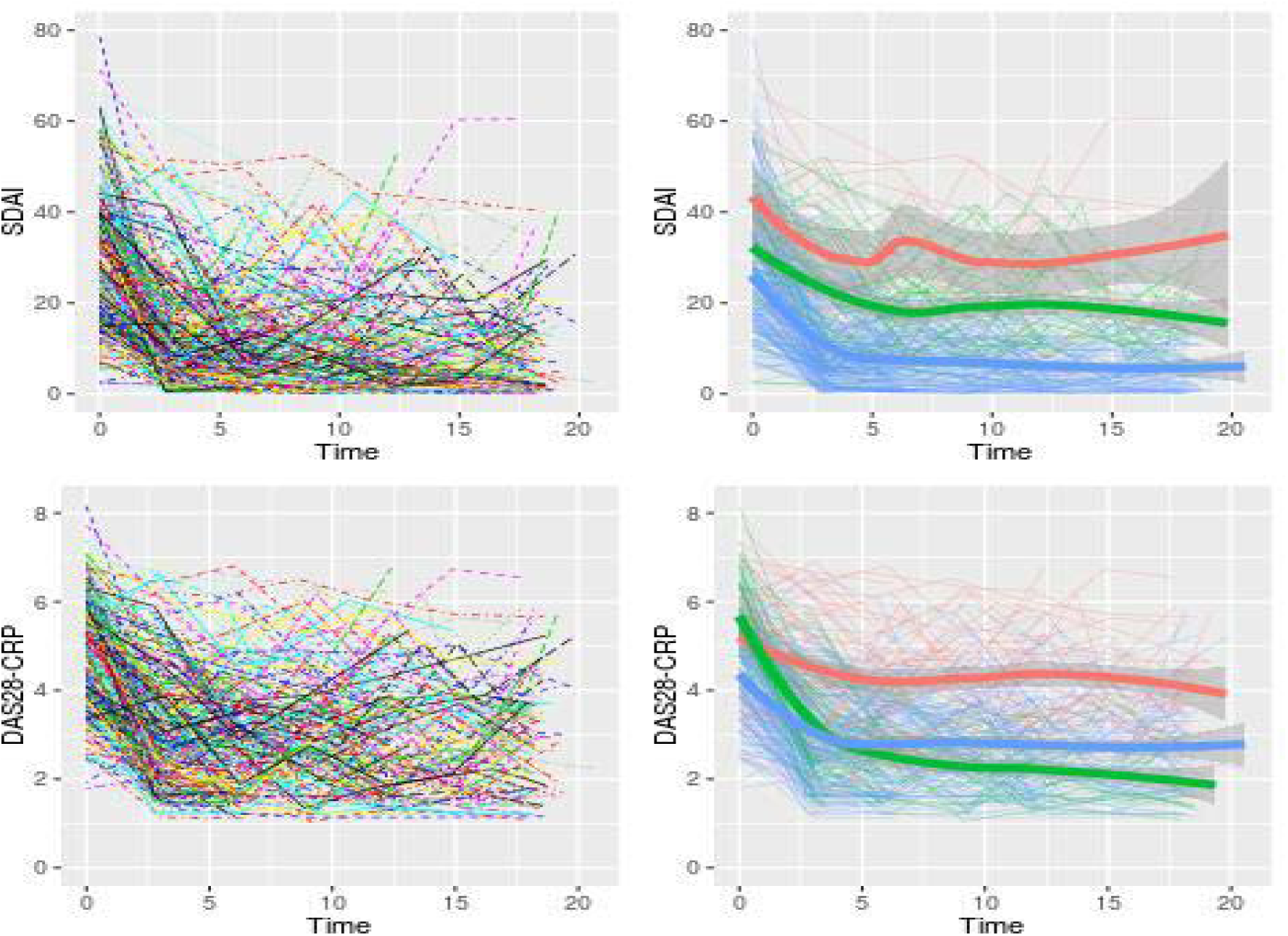
Individual and mean disease activity profiles over 18 months stratified by predicted class membership. *Based on SDAI, Class1 (Red; Inadequate Response [IR]): 17 (6.5%), Class 2 (Green; Higher Baseline Activity Responder [HBAR]): 59 (22.4%), Class 3 (Blue; Lower Baseline Activity Responder [LBAR]): 187 (71.1%). Based on DAS28-CRP, Class1 (Red; IR): 57 (21.7%), Class 2 (Green; HBAR): 56 (21.3%), Class 3 (Blue; LBAR): 150 (57.0%).

Table 4 and Supplementary Table 2 show the results of fitting latent class mixed models to SDAI and DAS28-CRP respectively when accounting for the baseline prescribing of medication. Both models provide evidence for three distinct sub-populations or latent classes. Class 1 corresponds to an **Inadequate Responder (IR)** group that, on average, started with high baseline disease activity that early on showed improvement, in association with initial medication, and then plateaued with moderate to high levels of disease activity. Class 2 corresponds to a **Higher Baseline Activity Responder (HBAR)** group that, on average, started with high baseline disease activity but showed sustained improvement over time. Class 3 corresponds to a **Lower Baseline Activity Responder (LBAR)** group that, on average, started with moderate levels of disease activity and showed sustained improvement. (See right hand side plots of Figure 1 and Table 5.)

**Table 4:**
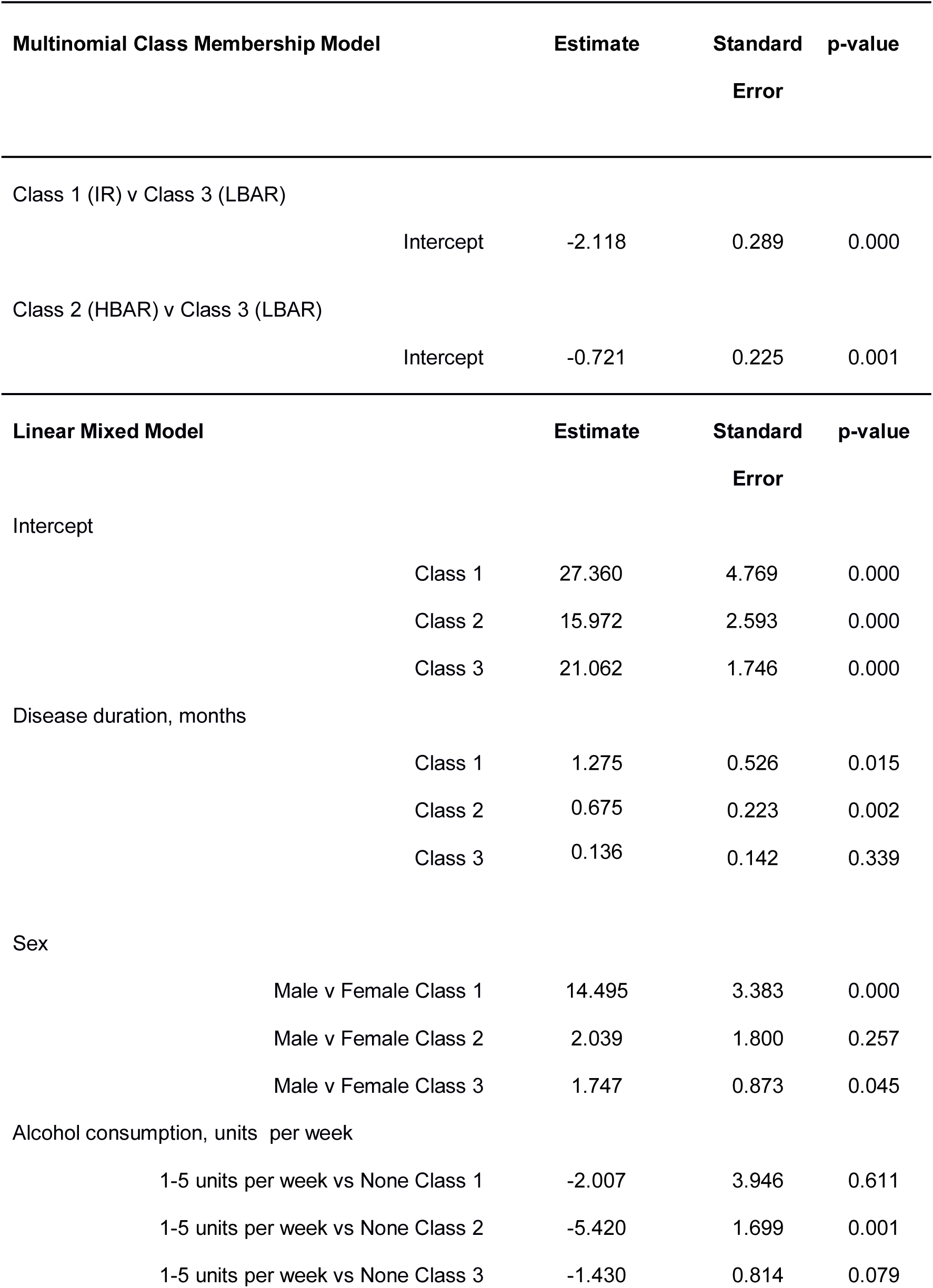
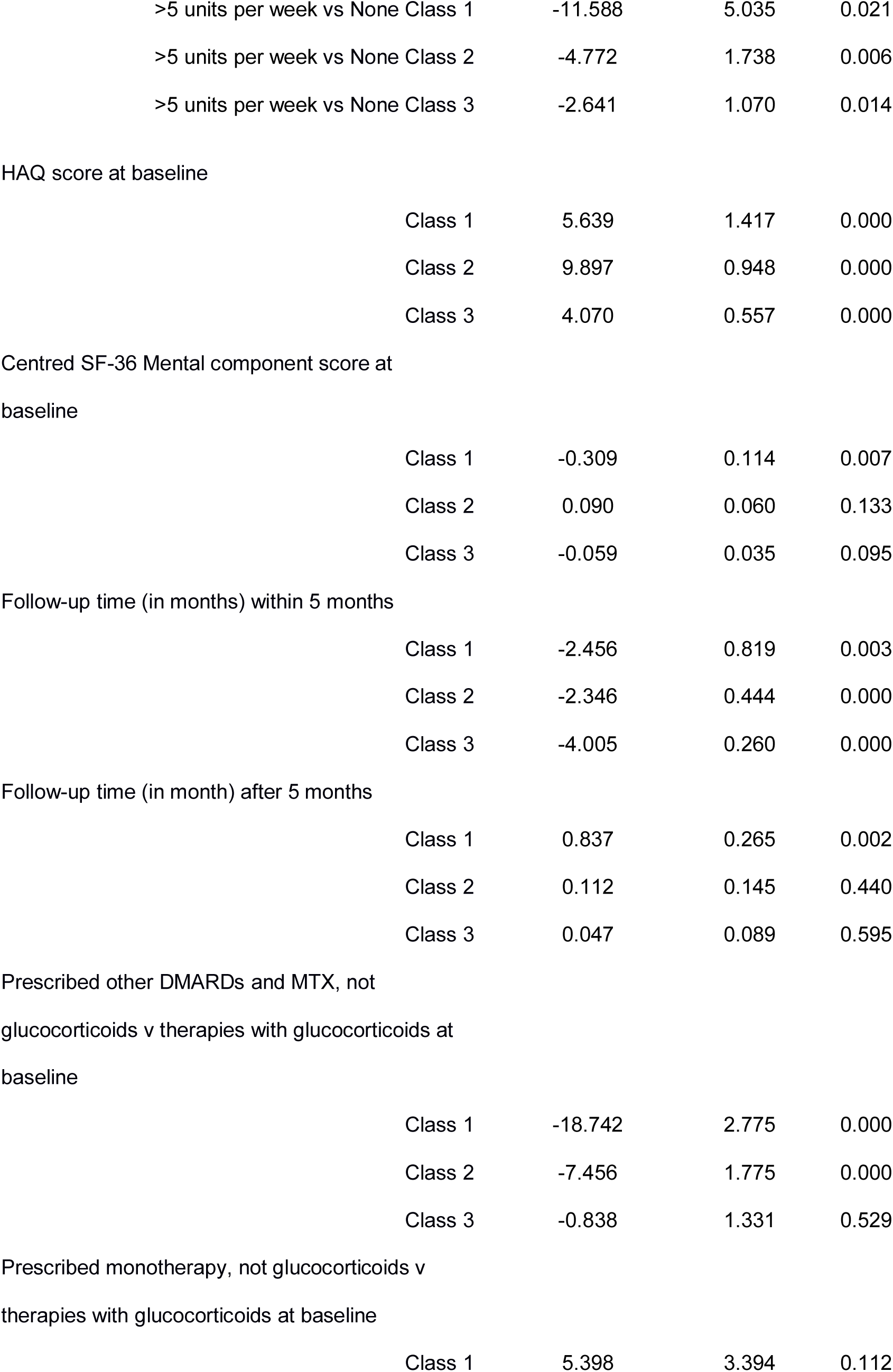
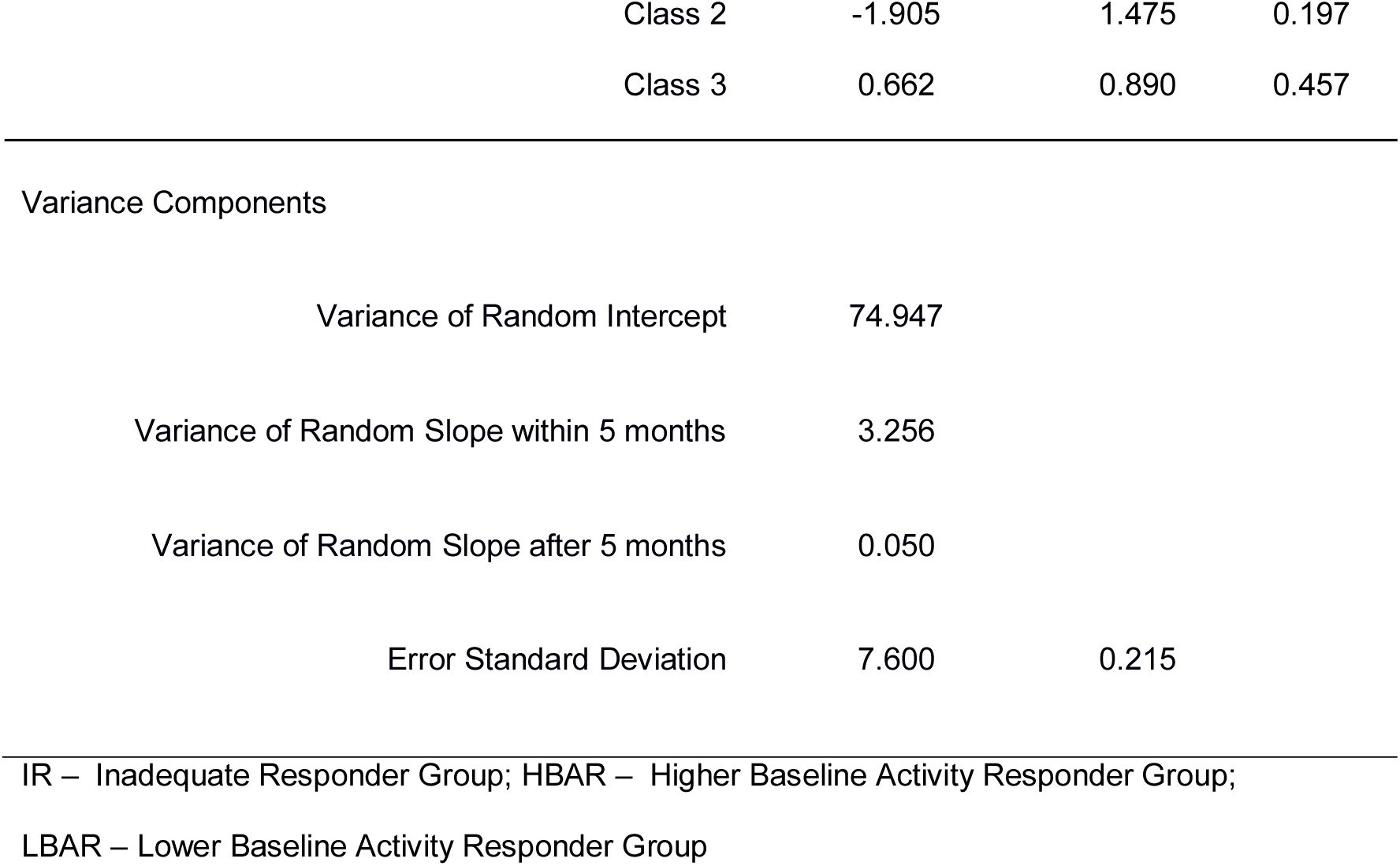
Latent Class Mixed Model for SDAI while Controlling for Baseline Medication: 3 Classes

| **Multinomial Class Membership Model** |  | **Estimate** | **Standard Error** | **p-value** |
| --- | --- | --- | --- | --- |
| Class 1 (IR) v Class 3 (LBAR) |  |  |  |  |
|  | Intercept | -2.118 | 0.289 | 0.000 |
| Class 2 (HBAR) v Class 3 (LBAR) |  |  |  |  |
|  | Intercept | -0.721 | 0.225 | 0.001 |
| **Linear Mixed Model** |  | **Estimate** | **Standard Error** | **p-value** |
| Intercept |  |  |  |  |
|  | Class 1 | 27.360 | 4.769 | 0.000 |
|  | Class 2 | 15.972 | 2.593 | 0.000 |
|  | Class 3 | 21.062 | 1.746 | 0.000 |
| Disease duration, months |  |  |  |  |
|  | Class 1 | 1.275 | 0.526 | 0.015 |
|  | Class 2 | 0.675 | 0.223 | 0.002 |
|  | Class 3 | 0.136 | 0.142 | 0.339 |
| Sex |  |  |  |  |
|  | Male v Female Class 1 | 14.495 | 3.383 | 0.000 |
|  | Male v Female Class 2 | 2.039 | 1.800 | 0.257 |
|  | Male v Female Class 3 | 1.747 | 0.873 | 0.045 |
| Alcohol consumption, units per week |  |  |  |  |
|  | 1-5 units per week vs None Class 1 | -2.007 | 3.946 | 0.611 |
|  | 1-5 units per week vs None Class 2 | -5.420 | 1.699 | 0.001 |
|  | 1-5 units per week vs None Class 3 | -1.430 | 0.814 | 0.079 |

|  |  |  |  |
| --- | --- | --- | --- |
| >5 units per week vs None Class 1 | -11.588 | 5.035 | 0.021 |
| >5 units per week vs None Class 2 | -4.772 | 1.738 | 0.006 |
| >5 units per week vs None Class 3 | -2.641 | 1.070 | 0.014 |
| HAQ score at baseline |  |  |  |
| Class 1 | 5.639 | 1.417 | 0.000 |
| Class 2 | 9.897 | 0.948 | 0.000 |
| Class 3 | 4.070 | 0.557 | 0.000 |
| Centred SF-36 Mental component score at baseline |  |  |  |
| Class 1 | -0.309 | 0.114 | 0.007 |
| Class 2 | 0.090 | 0.060 | 0.133 |
| Class 3 | -0.059 | 0.035 | 0.095 |
| Follow-up time (in months) within 5 months |  |  |  |
| Class 1 | -2.456 | 0.819 | 0.003 |
| Class 2 | -2.346 | 0.444 | 0.000 |
| Class 3 | -4.005 | 0.260 | 0.000 |
| Follow-up time (in month) after 5 months |  |  |  |
| Class 1 | 0.837 | 0.265 | 0.002 |
| Class 2 | 0.112 | 0.145 | 0.440 |
| Class 3 | 0.047 | 0.089 | 0.595 |
| Prescribed other DMARDs and MTX, not glucocorticoids v therapies with glucocorticoids at baseline |  |  |  |
| Class 1 | -18.742 | 2.775 | 0.000 |
| Class 2 | -7.456 | 1.775 | 0.000 |
| Class 3 | -0.838 | 1.331 | 0.529 |
| Prescribed monotherapy, not glucocorticoids v therapies with glucocorticoids at baseline |  |  |  |
| Class 1 | 5.398 | 3.394 | 0.112 |

|  |  |  |  |
| --- | --- | --- | --- |
| Class 2 | -1.905 | 1.475 | 0.197 |
| Class 3 | 0.662 | 0.890 | 0.457 |

Variance Components
|  |  |  |
| --- | --- | --- |
| Variance of Random Intercept | 74.947 |  |
| Variance of Random Slope within 5 months | 3.256 |  |
| Variance of Random Slope after 5 months | 0.050 |  |
| Error Standard Deviation | 7.600 | 0.215 |

IR – Inadequate Responder Group; HBAR – Higher Baseline Activity Responder Group; LBAR – Lower Baseline Activity Responder Group

**Table 5:**
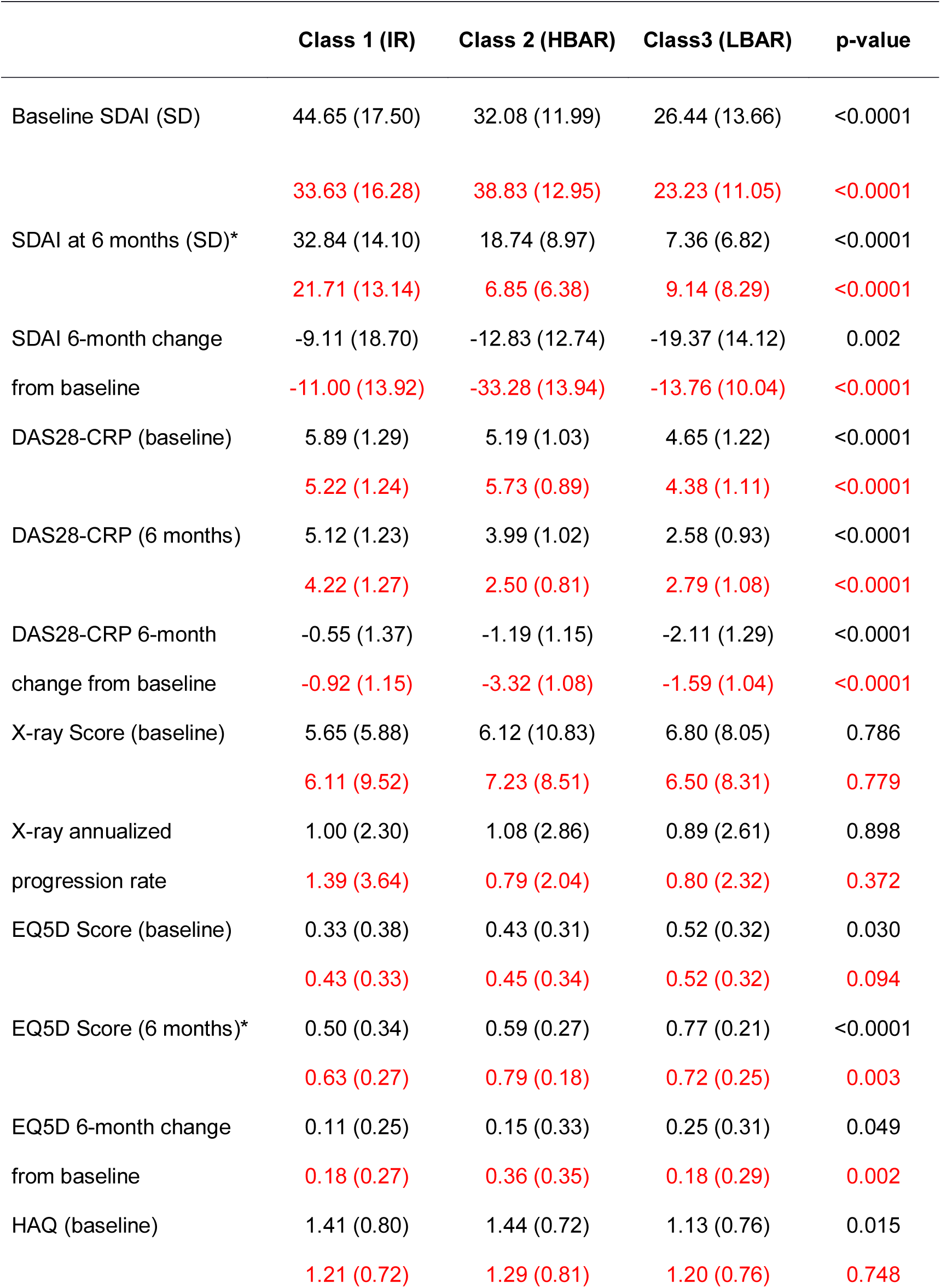
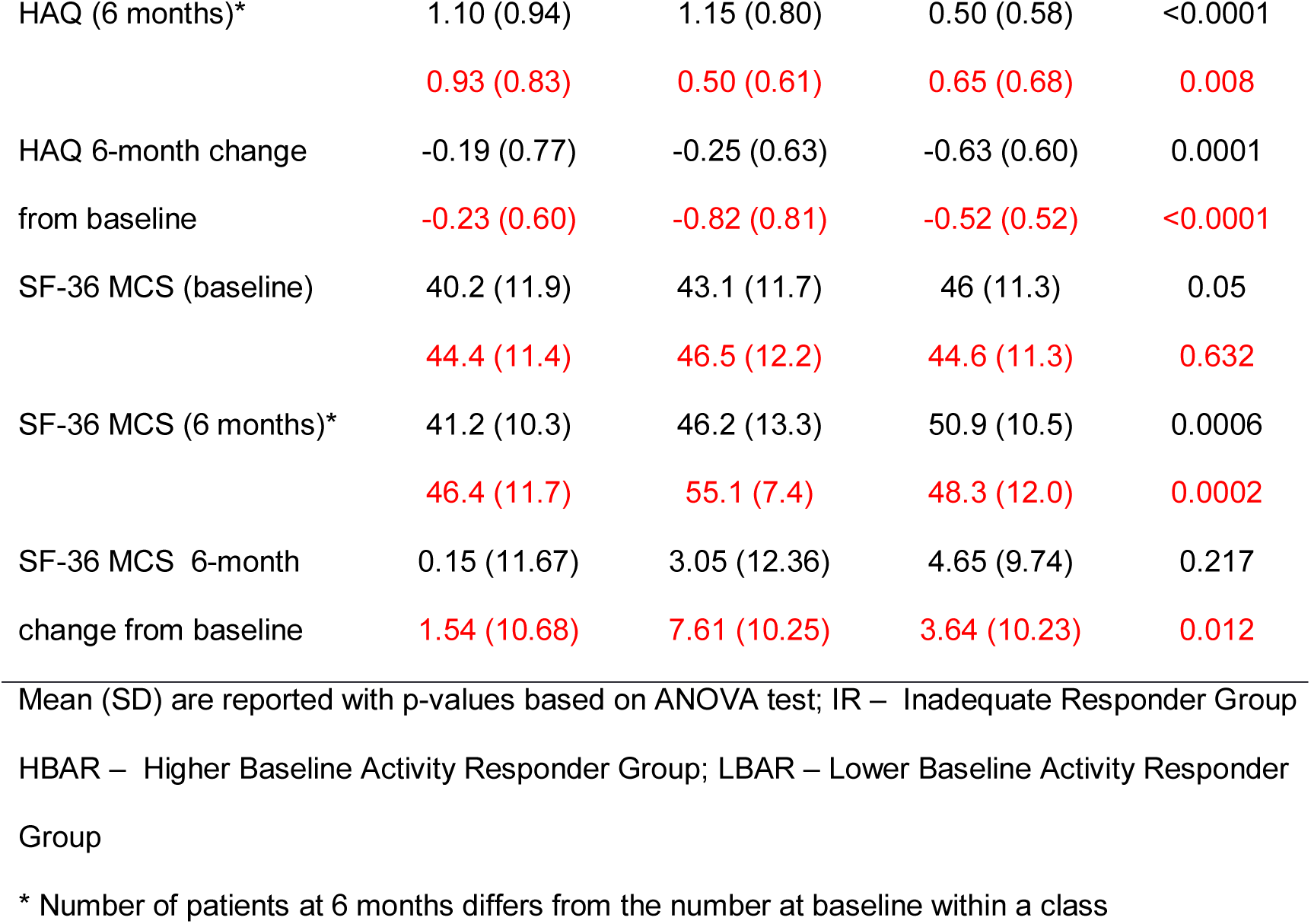
Disease outcomes by allocated SDAI (1^st^ row of cell) and DAS28-CRP (2^nd^ row of cell) classes, controlling for baseline medication

|  | Class 1 (IR) | Class 2 (HBAR) | Class3 (LBAR) | p-value |
| --- | --- | --- | --- | --- |
| Baseline SDAI (SD) | 44.65 (17.50) | 32.08 (11.99) | 26.44 (13.66) | <0.0001 |
|  | 33.63 (16.28) | 38.83 (12.95) | 23.23 (11.05) | <0.0001 |
| SDAI at 6 months (SD)* | 32.84 (14.10) | 18.74 (8.97) | 7.36 (6.82) | <0.0001 |
|  | 21.71 (13.14) | 6.85 (6.38) | 9.14 (8.29) | <0.0001 |
| SDAI 6-month change | -9.11 (18.70) | -12.83 (12.74) | -19.37 (14.12) | 0.002 |
| from baseline | -11.00 (13.92) | -33.28 (13.94) | -13.76 (10.04) | <0.0001 |
| DAS28-CRP (baseline) | 5.89 (1.29) | 5.19 (1.03) | 4.65 (1.22) | <0.0001 |
|  | 5.22 (1.24) | 5.73 (0.89) | 4.38 (1.11) | <0.0001 |
| DAS28-CRP (6 months) | 5.12 (1.23) | 3.99 (1.02) | 2.58 (0.93) | <0.0001 |
|  | 4.22 (1.27) | 2.50 (0.81) | 2.79 (1.08) | <0.0001 |
| DAS28-CRP 6-month | -0.55 (1.37) | -1.19 (1.15) | -2.11 (1.29) | <0.0001 |
| change from baseline | -0.92 (1.15) | -3.32 (1.08) | -1.59 (1.04) | <0.0001 |
| X-ray Score (baseline) | 5.65 (5.88) | 6.12 (10.83) | 6.80 (8.05) | 0.786 |
|  | 6.11 (9.52) | 7.23 (8.51) | 6.50 (8.31) | 0.779 |
| X-ray annualized | 1.00 (2.30) | 1.08 (2.86) | 0.89 (2.61) | 0.898 |
| progression rate | 1.39 (3.64) | 0.79 (2.04) | 0.80 (2.32) | 0.372 |
| EQ5D Score (baseline) | 0.33 (0.38) | 0.43 (0.31) | 0.52 (0.32) | 0.030 |
|  | 0.43 (0.33) | 0.45 (0.34) | 0.52 (0.32) | 0.094 |
| EQ5D Score (6 months)* | 0.50 (0.34) | 0.59 (0.27) | 0.77 (0.21) | <0.0001 |
|  | 0.63 (0.27) | 0.79 (0.18) | 0.72 (0.25) | 0.003 |
| EQ5D 6-month change | 0.11 (0.25) | 0.15 (0.33) | 0.25 (0.31) | 0.049 |
| from baseline | 0.18 (0.27) | 0.36 (0.35) | 0.18 (0.29) | 0.002 |
| HAQ (baseline) | 1.41 (0.80) | 1.44 (0.72) | 1.13 (0.76) | 0.015 |
|  | 1.21 (0.72) | 1.29 (0.81) | 1.20 (0.76) | 0.748 |
| HAQ (6 months)* | 1.10 (0.94) | 1.15 (0.80) | 0.50 (0.58) | <0.0001 |
|  | 0.93 (0.83) | 0.50 (0.61) | 0.65 (0.68) | 0.008 |
| HAQ 6-month change | -0.19 (0.77) | -0.25 (0.63) | -0.63 (0.60) | 0.0001 |
| from baseline | -0.23 (0.60) | -0.82 (0.81) | -0.52 (0.52) | <0.0001 |
| SF-36 MCS (baseline) | 40.2 (11.9) | 43.1 (11.7) | 46 (11.3) | 0.05 |
|  | 44.4 (11.4) | 46.5 (12.2) | 44.6 (11.3) | 0.632 |
| SF-36 MCS (6 months)* | 41.2 (10.3) | 46.2 (13.3) | 50.9 (10.5) | 0.0006 |
|  | 46.4 (11.7) | 55.1 (7.4) | 48.3 (12.0) | 0.0002 |
| SF-36 MCS 6-month | 0.15 (11.67) | 3.05 (12.36) | 4.65 (9.74) | 0.217 |
| change from baseline | 1.54 (10.68) | 7.61 (10.25) | 3.64 (10.23) | 0.012 |

Mean (SD) are reported with p-values based on ANOVA test; IR – Inadequate Responder Group
HBAR – Higher Baseline Activity Responder Group; LBAR – Lower Baseline Activity Responder
Group
\* Number of patients at 6 months differs from the number at baseline within a class

2

The IR, HBAR and LBAR groups were estimated to comprise 6.5%, 22.4% and 71.1% of the patients based on the SDAI model and 21.7%, 21.3% and 57% of the patients based on the DAS28-CRP model. No overall statistically significant differences in mean age at entry, disease duration, SF-36 physical component score at entry, sex, smoking status, alcohol consumption and serology distributions were found across latent classes. However mean BMI (p=0.064), mean baseline levels of functional disability (p=0.015) and SF-36 mental component scores (p=0.05) either were close to being or were statistically significantly different across the three classes/groups based on the SDAI model but not DAS28-CRP model. Table 5 shows that, based on the SDAI model, the LBAR group had on average significantly less functional disability than the other groups. A similar pattern was seen for mental health, with, on average, better mental health scores seen in the LBAR group (mean of 46) compared to both IR and HBAR (means of 40.2 and 43.1 respectively). Mean BMI was approximately 3.5kg/m higher in the IR group compared to the other SDAI groups. No evidence was found that prescribing behaviour of clinicians varied across the SDAI or DAS28-CRP defined groups. (See Supplementary Table 3.)

In the **IR** group, lower disease activity levels over time (measured by SDAI) were associated with being female, shorter symptom duration, consuming greater than 5 units of alcohol per week, less functional disability and better mental well-being at baseline. Being prescribed dual therapy of MTX and a second DMARD at baseline was associated with lower SDAI over time compared to either receiving monotherapy (excluding glucocorticoids) or therapies that included glucocorticoid usage (see Table 4).

For the **HBAR** group, lower levels of SDAI over time were associated with shorter symptom duration, increasing levels of alcohol consumption, and less functional disability. Moreover lower levels of disease activity were associated with being prescribed dual therapy of MTX and a second DMARD at baseline. In the **LBAR** group, being female, consuming alcohol and lower level of functional disability at baseline were associated with lower levels of SDAI.

Based on the DAS28-CRP model (Supplementary Table 2), lower levels of functional disability, better mental well-being and dual therapy of MTX with a second DMARD at baseline were associated with lower DAS28-CRP over time in the IR group. Higher levels of functional disability and receiving monotherapy (excluding glucocorticoids) were associated with higher levels of disease activity in the HBAR group. Whilst being female, drinking alcohol, having lower levels of functional disability and better mental health were associated with lower DAS28-CRP over time in the LBAR group.

Table 5 summarises other patient outcome data (X-ray score, EQ-5D, HAQ, SF-36 MCS) by allocated latent trajectory class. Class assignment could be made for 263 of the 267 eligible patients with some baseline and outcome information. The average EQ-5D scores both at baseline and 6 months were significantly higher in the LBAR group than the other groups as defined by the SDAI model (p<0.0001 and 0.049), whilst average level of functional disability remained significantly lower in this group at 6 months (p<0.0001). There was no statistical evidence for a difference in average X-ray annualised progression rate across groups.

## DISCUSSION

The TACERA cohort provides a unique opportunity to follow newly diagnosed and initially medication naïve patients in the UK, whose clinical outcomes are linked to extensive and detailed biological phenotyping (to be reported separately). We focused solely on seropositive RA patients whom have few non-severe comorbidities to reduce heterogeneity and to identify factors associated with outcomes in this subset.

We found clear evidence that remission at 6 months, whether defined using SDAI or DAS28-CRP, was associated with lower levels of disease activity and functional disability at baseline and with alcohol consumption at baseline. Aletaha et al.(27) have previously shown that disease activity early in the course of treatment predicts response to therapy after 1 year. However, other variables previously identified as associated with remission, such as age, gender, ethnicity and RA medications were not found to be statistically significant in our study. This may be due to the power of the current study (especially when restricted to the outcome defined at a single time point), to the use of a treat-to-target medication strategy, or to the fact that some variables may be less important in a pure seropositive disease cohort.

We also identified three trajectories of disease course and potential factors associated with them and also with disease activity levels within trajectories. We used the same trajectory labels (IR, HBAR and LBAR) in both SDAI and DAS28-CRP models, but the estimated proportion of patients who fell into the three groups varied according to the way disease activity was measured. Moreover the agreement between these two classifications was weak (Cohen’s Kappa of 0.18); and significant overall associations of baseline functional disability and SF-36 mental component score and near significant association with BMI were found only with the SDAI trajectories; thus suggesting these disease activity indices are not interchangeable.

Interestingly, we found no evidence for differences in prescribing behaviour at baseline amongst the trajectory classes irrespective of how trajectory classifications were made. However we found clear evidence that, even as early as 6 months, disease outcomes (e.g. EQ-5D, HAQ) and their changes from baseline differed between trajectory groups, with the IR group generally having the least improvement and poorer health outcomes. No evidence for differing annualized X-ray progression rates were seen across groups. This may be expected given the short length of follow-up and early intensive management. The findings of very few if any significant associations between standard clinical measures and trajectory classes, and evidence of clear changes in outcomes as early as 6 months would possibly suggest that these trajectories may instead be linked to distinct immunopathogenic subsets. Thus immunological biomarkers at baseline and early in follow-up (before 6 months) may be useful in identifying patients who beyond 6 months will have inadequate response to initial synthetic disease modifying treatment. We are currently investigating these trajectories from a biological viewpoint, utilising the extensive biological dataset collected in TACERA.

Heterogeneity in the effects of covariates on average SDAI level over time was seen across trajectories. Specifically, we found that effect sizes differed across trajectories with respect to functional disability, mental health, alcohol consumption and combination MTX with second DMARD compared to therapies with glucocorticoid usage. This provides evidence to support a stratified approach to management and treatment of patients falling into different trajectory groups.

Our results concerning the effect of alcohol consumption on both SDAI remission at 6 months and disease activity levels within SDAI trajectories suggest that consuming alcohol is associated with higher remission rates and lower disease activity over time. Similar inverse associations have been reported by other studies.(28–32) These findings may be due to alcohol intake being associated with lower levels of inflammatory markers and possibly because certain alcoholic drinks, for example red wine, may be anti-oxidant due to their flavonoid content, counteracting postprandial oxidative stress.(33,34)

The findings that mental health is a predictor of disease activity over time in the IR group and also possibly the LBAR group but not the HBAR group are worth highlighting. One-year results from the Scottish Early Rheumatoid Arthritis Inception (SERA) cohort have identified that predictors of functional disability at 1 year appear to be dominated by psychosocial rather than more traditional clinical measures, emphasising the potential benefit which may result from early access to non- pharmacological interventions targeting key psychosocial factors, such as mental health and work disability.(35) The possibility of adopting a non-pharmacological intervention strategy for those in the IR and LBAR (that make up approximately 78% of the RA patients) should be explored in more depth.

Latent class mixed modelling approaches have only recently begun to be used to investigate whether distinct subpopulations exist within RA with respect to disease course.(5,36,37) Previous studies using DAS28 have identified similar trajectory groups. The larger study by Barnabe et al. was able to identify five trajectory groups instead of three. However combining their Groups 1 and 4 (i.e. both with initial high activity) into a single group would coincide with our HBAR group, whilst combining their Groups 2 and 3 (i.e. both with initial moderate activity) would correspond to our LBAR group. Our work applies also to SDAI and demonstrates similar trajectory groups within a purely seropositive population. Additionally, we controlled for factors that may impact on disease activity over time within these trajectory groups.

## CONCLUSION

In conclusion, we found that, in early seropositive RA with few non-severe comorbidities, lower baseline levels of functional disability and disease activity, along with alcohol intake, are associated with 6-month clinical remission. We identified three subpopulations based on disease trajectories that not only differ in terms of disease course but also with regard to the effect of risk factors, such as mental well-being, on disease activity over time. Our data further highlight RA heterogeneity (i.e. trajectory classes) not explainable by clinical factors and indicate the possible use of biomarkers collected at baseline and early follow-up to help patient management and better targeting of existing and novel therapies.

## Abbreviations

### Availability of data and materials

Request for access to RA-MAP data and samples is

## Authors’ contribution and competing interests

A list of contributing authors with disclosure of possible conflicts of interests can be found in “RA- MAP Contributing Authors_ART_2019.docx”

## Data Availability

Data are available through the RA-MAP Sample and Data Access Committee. Contact: Dr Dennis Lendrem

## Acknowledgements

We acknowledge the support of TACERA Principal Investigators from all contributing NHS sites and the members of the TACERA Study Steering and Data Monitoring Committee. The Research was supported by the NIHR Newcastle Biomedical Research Centre. Members of the RA-MAP Consortium can be found in the document “RA-MAP Consortium Membership_ART_2019.docx”.

## Ethics approval and consent to participate

Ethical approval was authorised by the National Research Ethics Service London Central Committee (Reference number: 12/LO/0469). Informed, written consent was obtained.

## Consent for publication

Not applicable

## REFERENCE

1. Scott DL, Wolfe F. Huizinga TW. Rheum arthritis Lancet. 2010;376:1094–108.

2. Cook MJ, Diffin J, Scire CA, Lunt M, MacGregor AJ, Symmons DPM, et al. Predictors and outcomes of sustained, intermittent or never achieving remission in patients with recent onset inflammatory polyarthritis: results from the Norfolk Arthritis Register. Rheumatology. 2016;55(9):1601–9.

3. Ma MHY, Ibrahim F, Kingsley GH, Cope A, Scott DL. Variable impacts of different remission states on health-related quality of life in rheumatoid arthritis. Clin Exp Rheumatol. 2018;36(2):203–9.

4. Wolfe F, Rasker JJ, Boers M, Wells GA, Michaud K. Minimal disease activity, remission, and the long-term outcomes of rheumatoid arthritis. Arthritis Care Res (Hoboken). 2007;57(6):935–42.

5. Siemons L, ten Klooster PM, Vonkeman HE, Glas CAW, van de Laar MAFJ. Distinct trajectories of disease activity over the first year in early rheumatoid arthritis patients following a treat-to-target strategy. Arthritis Care Res (Hoboken). 2014;66(4):625–30.

6. Norton S, Fu B, Scott DL, Deighton C, Symmons DPM, Wailoo AJ, et al. Health Assessment Questionnaire disability progression in early rheumatoid arthritis: systematic review and analysis of two inception cohorts. In: Seminars in arthritis and rheumatism. 2014. p. 131–44.

7. Bukhari M, Thomson W, Naseem H, Bunn D, Silman A, Symmons D, et al. The performance of anti--cyclic citrullinated peptide antibodies in predicting the severity of radiologic damage in inflammatory polyarthritis: results from the Norfolk Arthritis Register. Arthritis Rheum Off J Am Coll Rheumatol. 2007;56(9):2929–35.

8. Van Gaalen FA, Van Aken J, Huizinga TWJ, Schreuder GMT, Breedveld FC, Zanelli E, et al. Association between HLA class II genes and autoantibodies to cyclic citrullinated peptides (CCPs) influences the severity of rheumatoid arthritis. Arthritis Rheum Off J Am Coll Rheumatol. 2004;50(7):2113–21.

9. Cope AP, Barnes MR, Belson A, Binks M, Brockbank S, Bonachela-Capdevila F, et al. The RA-MAP Consortium: a working model for academia--industry collaboration. Nat Rev Rheumatol. 2018;14(1):53.

10. Aletaha D, Neogi T, Silman AJ, Funovits J, Felson DT, Bingham III CO, et al. 2010 rheumatoid arthritis classification criteria: an American College of Rheumatology/European League Against Rheumatism collaborative initiative. Arthritis Rheum. 2010;62(9):2569–81.

11. Arnett FC, Edworthy SM, Bloch DA, Mcshane DJ, Fries JF, Cooper NS, et al. The American Rheumatism Association 1987 revised criteria for the classification of rheumatoid arthritis. Arthritis Rheum Off J Am Coll Rheumatol. 1988;31(3):315–24.

12. NICE. The management of rheumatoid arthritis in adults. (Clinical guideline 79.) [Internet]. London; 2009. Available from: www.nice.org.uk/CG79

13. Aletaha D, Smolen J. The Simplified Disease Activity Index (SDAI) and the Clinical Disease Activity Index (CDAI): a review of their usefulness and validity in rheumatoid arthritis. Clin Exp Rheumatol. 2005;23(5):S100.

14. Anderson J, Caplan L, Yazdany J, Robbins ML, Neogi T, Michaud K, et al. Rheumatoid arthritis disease activity measures: American College of Rheumatology recommendations for use in clinical practice. Arthritis Care Res (Hoboken). 2012;64(5):640–7.

15. Anderson JK, Zimmerman L, Caplan L, Michaud K. Measures of rheumatoid arthritis disease activity: Patient (PtGA) and Provider (PrGA) Global Assessment of Disease Activity, Disease Activity Score (DAS) and Disease Activity Score with 28-Joint Counts (DAS28), Simplified Disease Activity Index (SDAI), Clinical Disease Activity Index (CDAI), Patient Activity Score (PAS) and Patient Activity Score-II (PASII), Routine Assessment of Patient Index Data (RAPID), Rheumatoid Arthritis Disease Activity Index (RADAI) and Rheumatoid Arthritis Disease Activity Index-5 (RADAI-5), Chronic Arthritis Systemic Index (CASI), Patient-Based Disease Activity Score With ESR (PDAS1) and Patient-Based Disease Activity Score without ESR (PDAS2), and Mean Overall Index for Rheumatoid Arthritis (MOI-RA). Arthritis Care Res (Hoboken). 2011;63(S11):S14–S36.

16. Dougados M, Aletaha D, van Riel P. Disease activity measures for rheumatoid arthritis. Clin Exp Rheumatol. 2007;25(5):S22.

17. Felson DT, Smolen JS, Wells G, Zhang B, Van Tuyl LHD, Funovits J, et al. American College of Rheumatology/European League Against Rheumatism provisional definition of remission in rheumatoid arthritis for clinical trials. Arthritis Rheum. 2011;63(3):573–86.

18. Larsen A. A radiological method for grading the severity of rheumatoid arthritis. Scand J Rheumatol. 1975;4(4):225–33.

19. Larsen A, Dale K, Eek M. Radiographic evaluation of rheumatoid arthritis and related conditions by standard reference films. Acta Radiol Diagnosis. 1977;18(4):481–91.

20. Scott DL, Houssien DA, Laasonen L. Proposed modification to Larsen’s scoring methods for hand and wrist radiographs. Rheumatology. 1995;34(1):56.

21. Fries JF, Spitz P, Kraines RG, Holman HR. Measurement of patient outcome in arthritis. Arthritis Rheum. 1980;23(2):137–45.

22. Hurst NP, Kind P, Ruta D, Hunter M, Stubbings A. Measuring health-related quality of life in rheumatoid arthritis: validity, responsiveness and reliability of EuroQol (EQ-5D). Br J Rheumatol. 1997;36(5):551–9.

23. Linde L, Sørensen J, Østergaard M, Hørslev-Petersen K, Hetland ML. Health-related quality of life: validity, reliability, and responsiveness of SF-36, EQ-15D, EQ-5D, RAQoL, and HAQ in patients with rheumatoid arthritis. J Rheumatol. 2008;35(8):1528–37.

24. Ruta DA, Hurst NP, Kind P, Hunter M, Stubbings A. Measuring health status in British patients with rheumatoid arthritis: reliability, validity and responsiveness of the short form 36-item health survey (SF-36). Br J Rheumatol. 1998;37(4):425–36.

25. Team RC. R: A language and environment for statistical computing [Computer software]. Vienna, Austria R Found Stat Comput. 2017;

26. Charlson ME, Pompei P, Ales KL, MacKenzie CR. A new method of classifying prognostic comorbidity in longitudinal studies: development and validation. J Chronic Dis. 1987;40(5):373–83.

27. Aletaha D, Funovits J, Keystone EC, Smolen JS. Disease activity early in the course of treatment predicts response to therapy after one year in rheumatoid arthritis patients. Arthritis Rheum Off J Am Coll Rheumatol. 2007;56(10):3226–35.

28. Dunlop DD, Semanik P, Song J, Manheim LM, Shih V, Chang RW. Risk factors for functional decline in older adults with arthritis. Arthritis Rheum. 2005;52(4):1274–82.

29. Kim S-K, Park S-H, Shin I-H, Choe J-Y. Anti-cyclic citrullinated peptide antibody, smoking, alcohol consumption, and disease duration as risk factors for extraarticular manifestations in Korean patients with rheumatoid arthritis. J Rheumatol. 2008;35(6):995–1001.

30. Maxwell JR, Gowers IR, Moore DJ, Wilson AG. Alcohol consumption is inversely associated with risk and severity of rheumatoid arthritis. Rheumatology. 2010;49(11):2140–6.

31. Nissen MJ, Gabay C, Scherer A, Finckh A, in Rheumatoid Arthritis SCQMP. The effect of alcohol on radiographic progression in rheumatoid arthritis. Arthritis Rheum. 2010;62(5):1265–72.

32. Papadopoulos NG, Alamanos Y, Voulgari P V, Epagelis EK, Tsifetaki N, Drosos AA. Does cigarette smoking influence disease expression, activity and severity in early rheumatoid arthritis patients? Clin Exp Rheumatol. 2005;23(6):861.

33. Covas MI, Gambert P, Fitó M, de la Torre R. Wine and oxidative stress: up-to-date evidence of the effects of moderate wine consumption on oxidative damage in humans. Atherosclerosis. 2010;208(2):297–304.

34. Lu B, Solomon DH, Costenbader KH, Keenan BT, Chibnik LB, Karlson EW. Alcohol consumption and markers of inflammation in women with preclinical rheumatoid arthritis. Arthritis Rheum. 2010;62(12):3554–9.

35. Kronisch C, McLernon DJ, Dale J, Paterson C, Ralston SH, Reid DM, et al. Brief Report: Predicting Functional Disability: One-Year Results From the Scottish Early Rheumatoid Arthritis Inception Cohort. Arthritis Rheumatol. 2016;68(7):1596–602.

36. Barnabe C, Sun Y, Boire G, Hitchon CA, Haraoui B, Thorne JC, et al. Heterogeneous disease trajectories explain variable radiographic, function and quality of life outcomes in the Canadian Early Arthritis Cohort (CATCH). PLoS One. 2015;10(8):e0135327.

37. Wabe N, Sorich MJ, Wechalekar MD, Cleland LG, McWilliams L, Lee A, et al. Characterising deviation from treat-to-target strategies for early rheumatoid arthritis: the first three years. Arthritis Res Ther. 2015;17(1):48.

38. Charlson ME, Charlson RE, Peterson JC, Marinopoulos SS, Briggs WM, Hollenberg JP. The Charlson comorbidity index is adapted to predict costs of chronic disease in primary care patients. J Clin Epidemiol. 2008;61(12):1234–40.

